# Recent trends in postoperative mortality after liver resection- A systemic review and metanalysis of studies published in last 5 years and metaregression of various factors affecting 90 days mortality

**DOI:** 10.1101/2021.03.26.21254407

**Authors:** Bhavin Vasavada, Hardik Patel

## Abstract

**Aim:** The aim of this systemic review and meta-analysis was to analyse 90 days mortality after liver resection, and also study various factors associated with mortality via univariate and multivariate metaregression.

**Methods:** PubMed, Cochrane library, Embase, google scholar, web of science with keywords like ‘liver resection”; “mortality”;” hepatectomy”. Weighted percentage 90 days mortalities were analysed. univariate metaregression was done by DerSimonian-Liard methods. Major hepatectomy, open surgery, cirrhotic livers, blood loss, hepatectomy for hepatocellular carcinoma, hepatectomy for colorectal liver metastasis were taken as moderators in metaregression analysis. We decided to enter all co-variants in multivariate model to look for mixed effects. Heterogeneity was assessed using the Higgins I^2^ test, with values of 25%, 50% and 75% indicating low, moderate and high degrees of heterogeneity. Cohort studies were assessed for bias using the Newcastle-Ottawa Scale to assess for the risk of bias. Publication bias was assessed using funnel plot. Funnel plot asymmetry was evaluated by Egger’s test.

**Results:** Total 29931 patients’ data who underwent liver resections for various etiologies were pooled from 41 studied included1257 patients died within 90 days post operatively. Weighted 90 days mortality was 3.6% (95% C.I 2.8% −4.4%). However, heterogeneity of the analysis was high with I^2^ 94.625%.(p<0.001). We analysed various covariates like major hepatectomy, Age of the patient, blood loss, open surgery, liver resections done for hepatocellular carcinoma or colorectal liver metastasis and cirrhotic liver to check for their association with heterogeneity in the analysis and hence 90 days mortality. On univariate metaregression analysis major hepatectomy (p<0.001), Open hepatectomy (p<0.001), blood loss (p=0.002) was associated with heterogeneity in the analysis and 90 days mortality. On multivariate metaregression Major hepatectomy(p=0.003) and Open surgery (p=0.012) was independently associated with higher 90 days mortality, and liver resection for colorectal liver metastasis was independently associated with lesser 90 days mortality (z= −4.11,p<0.01). Residual heterogeneity after all factor multivariate metaregression model was none (I^2^=0,Tau^2^=0, H^2^=1) and nonsignificant (p=0.49).

**Conclusion:** Major hepatectomy, open hepatectomy, and cirrhotic background is associated with higher mortality rates and colorectal liver metastasis is associated with lower peri operative mortality rates.

## Introduction

Liver resection is now established curative treatment for various malignant and benign liver pathology. Currently Hepatocellular carcinoma and colorectal liver metastasis is the most common indications for liver surgery as well as primary and secondary liver malignancies. [1,2].

Perioperative care and surgical techniques for liver resections have improved significantly in past few years, resulting in improved perioperative outcomes. However, despite improvements liver resections are associated with variable outcomes according to centres, countries and surgeons and mortality and morbidity rates after this demanding surgery are still high and variable. [3,4,5]. There is also lack of uniformity in the literature regarding description of post-operative mortality with some mention in hospital mortality, some 30 days mortality and some 90 days mortality.

Recently published study by Filmann et al.[6] suggested higher In hospital mortality rate across the Germany. However due to lack of data they also could not report actual 90 days mortality data and mentioned in hospital mortality rates. However, their findings raised concerns about under reporting of mortality data in scientific community.

Aim of this systemic review and meta-analysis was to analyse 90 days mortality after liver resection, and study various factors associated with mortality via univariate and multivariate metaregression.

## Material and Methods

The study was conducted according to the Preferred Reporting Items for Systematic Reviews and Meta-Analyses (PRISMA) statement and MOOSE guidelines. [7,8]. We conducted literature search as described by Gossen et al. [9]. PubMed, Cochrane library, Embase, google scholar, web of science with keywords like ‘liver resection”; “mortality”;” hepatectomy”. Two independent authors extracted the data (B.V and H.P). We evaluated last studies published in last 5 years to see recent trends in 90 days mortality. In case of disagreements decision reached on basis of discussions.

### Definitions

Major hepatectomy was defined as resection of more than or equal to 3 segments. Post-operative mortality was defined as any mortality within 90 days post-operative period.

### Statistical Analysis

The meta-analysis was done using Review Manager 5.4 and JASP Team (2020). JASP (Version 0.14.1)(University of Amsterdam). Weighted percentage 90 days mortalities with 95% confidence intervals were used. Univariate metaregression was done by DerSimonian-Liard methods. Major hepatectomy, open surgery, cirrhotic livers, blood loss, hepatectomy for hepatocellular carcinoma, hepatectomy for colorectal liver metastasis were taken as co-variates in metaregression analysis to study their association heterogeneity of the meta-analysis and 90 days mortality. Factors with p value less than 0.05 were entered in multivariate metaregression model and then we decided to check for residual heterogeneity, if residual heterogeneity is still significant, we decided to enter all co-variants in multivariate model to look for mixed effects. Heterogeneity was assessed using the Higgins I^2^ test, with values of 25%, 50% and 75% indicating low, moderate and high degrees of heterogeneity, respectively and assessed p value for significance of heterogeneity and tau^2^ and H^2^ value [10]. The random effects model was used in meta-analysis.

### Assessment of Bias

Cohort studies were assessed for bias using the Newcastle-Ottawa Scale to assess for the risk of bias [11,12] Publication bias was assessed using funnel plot. Funnel plot asymmetry was evaluated by Egger’s test.

### Inclusion and Exclusion criteria for studies

#### Inclusion criteria

· Studies with full texts
· Studies published in last 5 years.
· Studies mentioning 90 days mortality rates.
· Studies which evaluated liver resections for different etiologies.
· Studies published in last 5 years.
· English language studies.

#### Exclusion criteria

· Studies which were not fulfilling above criteria.
· Duplicate studies.
· Studies which included analysis of liver resection for single etiology.

## Results

### Data extraction, study characteristics and quality assessment

‘PUBMED’, ‘SCOPUS’, and ‘EMBASE’ databases were searched using keywords and the search strategy described above. Initially 4246 studies published in last 5 years were screened from above search strategy and 23 additional records were screened from references of above studies. After removing duplicates 3343 studies screened again. 3265 studies excluded meeting the exclusion criteria. 78 full text articles were evaluated. 36 full texts article removed as they did not mention 90 days mortality and instead mentioned in hospital, perioperative or 30 days post-operative mortality.41 studies included in final qualitative and quantitative analysis. [Figure 1 Prisma flow chart]. Risk of bias summary is mentioned in Figure 2. Study characteristics are mentioned in table 1. All the studies included are either retrospective studies or propensity score matched analysis from retrospective data. In propensity score matched analysis unmatched total data were analysed for 90 days mortality, so effectively all the data were retrospective.

**Table 1:** Study Characteristics

| STUDY | MORTALITY | total | 90 day mortality (%) | Major HEPATECTOMY | Open surgery | Cirrhosis Present | Age (median) | blood loss (ml) | hepatocellular carcinoma (n) | Colorectal metastasis (n) |
| --- | --- | --- | --- | --- | --- | --- | --- | --- | --- | --- |
| al-saeedi2020 | 21 | 209 | 10 | 209 | 209 | 83 | 60 | 1500 | 16 | 59 |
| al-saif2019 | 5 | 111 | 4.5 | 48 | 89 |  | 52.6 |  | 14 | 1 |
| alam2016 | 4 | 77 | 5.2 | 45 | 77 |  | 49 |  | 11 | 35 |
| amico2016 | 6 | 84 | 7.1 | 38 | 84 | 6 | 54.01 |  | 9 | 30 |
| atili2019 | 1 | 10 | 10 | 10 | 8 | 0 | 60.3 |  | 3 | 0 |
| braunwarth2019 | 19 | 458 | 4.1 | 242 | 430 | 26 | 61 | 400 | 49 | 149 |
| chin2020 | 24 | 472 | 5.1 | 56 | 472 | 168 | 62 | 682 | 309 | 163 |
| CHOPINET2018 | 7 | 75 | 9.3 | 75 | 75 | 75 | 60 | 209 | 75 | 0 |
| CIESLAK2016 | 12 | 163 | 7.4 | 163 | 163 | 103 | 63 | 211 | 12 | 0 |
| DESARI2018 | 51 | 1269 | 4 | 645 | 1269 | 289 | 65.3 |  |  |  |
| DIGS2020 | 474 | 4263 | 11.1 | 4263 | 4263 |  | 60 |  |  |  |
| Durate2020 | 14 | 178 | 7.8 | 178 | 131 | 22 | 58 | 450 |  |  |
| fujii2019 | 6 | 103 | 5.8 | 103 | 103 | 63 | 67 | 2500 |  |  |
| garnier2018 | 5 | 111 | 4.5 | 111 | 111 | 37 | 66 |  | 9 | 66 |
| gelli2018 | 117 | 1803 | 6.5 | 619 | 1803 | 226 | 61 |  |  |  |
| gilg2016 | 138 | 4460 | 3.1 | 1615 |  |  | 64 |  | 393 | 2644 |
| Goh2017 | 1 | 195 | 0.5 | 12 | 0 | 50 | 60 | 200 | 102 | 46 |
| Grey2016 | 3 | 66 | 4.5 | 31 | 62 |  | 62 |  | 18 | 33 |
| gupta2018 | 0 | 26 | 1.9 | 12 | 26 |  | 82 | 400 | 4 | 20 |
| Goyri2019 | 17 | 701 | 2.4 | 348 |  |  | 63 |  | 76 | 491 |
| halls2017 | 16 | 2861 | 0.6 | 429 | 222 | 220 | 64 |  | 189 | 1630 |
| higashi2015 | 8 | 144 | 5.6 | 138 | 144 |  | 65.1 |  | 91 | 12 |
| hobeika2018 | 34 | 3150 | 1.1 | 80 | 0 | 774 | 62 | 350 | 1247 | 714 |
| **khaoudy2018** | 90 | 1906 | 4.7 | 1334 |  | 686 | 67.6 |  | 21 | 24 |
| **lee2020** | 0 | 72 | 0.7 | 72 |  |  | 57 |  | 48 | 9 |
| **lemke2017** | 39 | 749 | 5.2 | 516 | 681 | 25 | 64 |  | 56 | 489 |
| **Levisandri2018** | 9 | 225 | 4 | 225 |  |  | 62.4 | 400 | 77 | 106 |
| **Lin2019** | 3 | 281 | 1.1 |  | 232 |  | 55 | 300 |  |  |
| longbotthom2019 | 8 | 283 | 2.8 | 283 | 283 | 124 | 62 |  | 36 | 237 |
| longbotthom1.2019 | 4 | 49 | 8.2 | 49 | 49 | 20 | 78 |  |  |  |
| Matteo2017 | 2 | 340 | 0.6 | 48 | 340 |  | 65 | 300 | 82 | 196 |
| Ome2017 | 1 | 142 | 0.7 | 0 | 79 | 47 | 68 | 600 |  |  |
| Pietraz2018 | 5 | 195 | 2.6 | 47 | 0 |  | 68.2 |  | 22 | 130 |
| ratti2018 | 4 | 204 | 2 | 114 | 102 | 8 | 62 |  |  |  |
| ruzzenente2017 | 11 | 803 | 1.4 | 242 | 803 | 109 | 65.54 |  | 176 | 279 |
| saleema2017 | 5 | 75 | 6.7 | 30 | 75 | 30 | 52 | 665 | 37 | 10 |
| urbani2017 | 1 | 60 | 1.7 | 0 | 60 |  | 60 | 150 | 7 | 48 |
| vanmirelo2016 | 37 | 956 | 3.9 | 466 | 956 | 320 | 64 | 600 | 61 | 657 |
| vigano2020 | 50 | 2212 | 2.3 | 2212 | 2212 | 0 | 62 |  | 414 |  |
| worhunsky2016 | 2 | 167 | 1.2 | 30 | 0 | 48 | 58 | 100 | 56 |  |
| yip2016 | 3 | 223 | 1.3 | 103 | 200 |  | 67 |  | 25 |  |
| zarour2019 | 1 | 45 | 2.2 | 23 | 45 | 8 | 75 |  | 45 |  |

**Table 2:** Univariate and Multivariate meta regression analysis.

| Factors | Z value in<br>univariate<br>metaregression | P value in<br>univariate<br>metaregression | Z value in<br>multivariate<br>metaregression<br>(ALL<br>FACTORS) | P value in<br>multivariate<br>metaregression<br>(ALL<br>FACTORS) |
| --- | --- | --- | --- | --- |
| Major<br>Hepatectomy | **3.995** | **<0.001** | **2.945** | **0.003** |
| Open Surgery | **4.762** | **<0.001** | **2.504** | **0.012** |
| **Blood loss** | **3.073** | **0.002** | 0.762 | 0.446 |
| Age | -1.046 | 0.296 | -1.79 | 0.73 |
| Cirrhosis | -0.579 | 0.562 | **3.004** | **0.003** |
| Hepatocellular Carcinoma | -1.633 | 0.103 | -0.608 | 0.543 |
| Colorectal Liver Metastasis | -1.210 | 0.226 | **-4.116** | **<0.001** |

**Figure 1.**
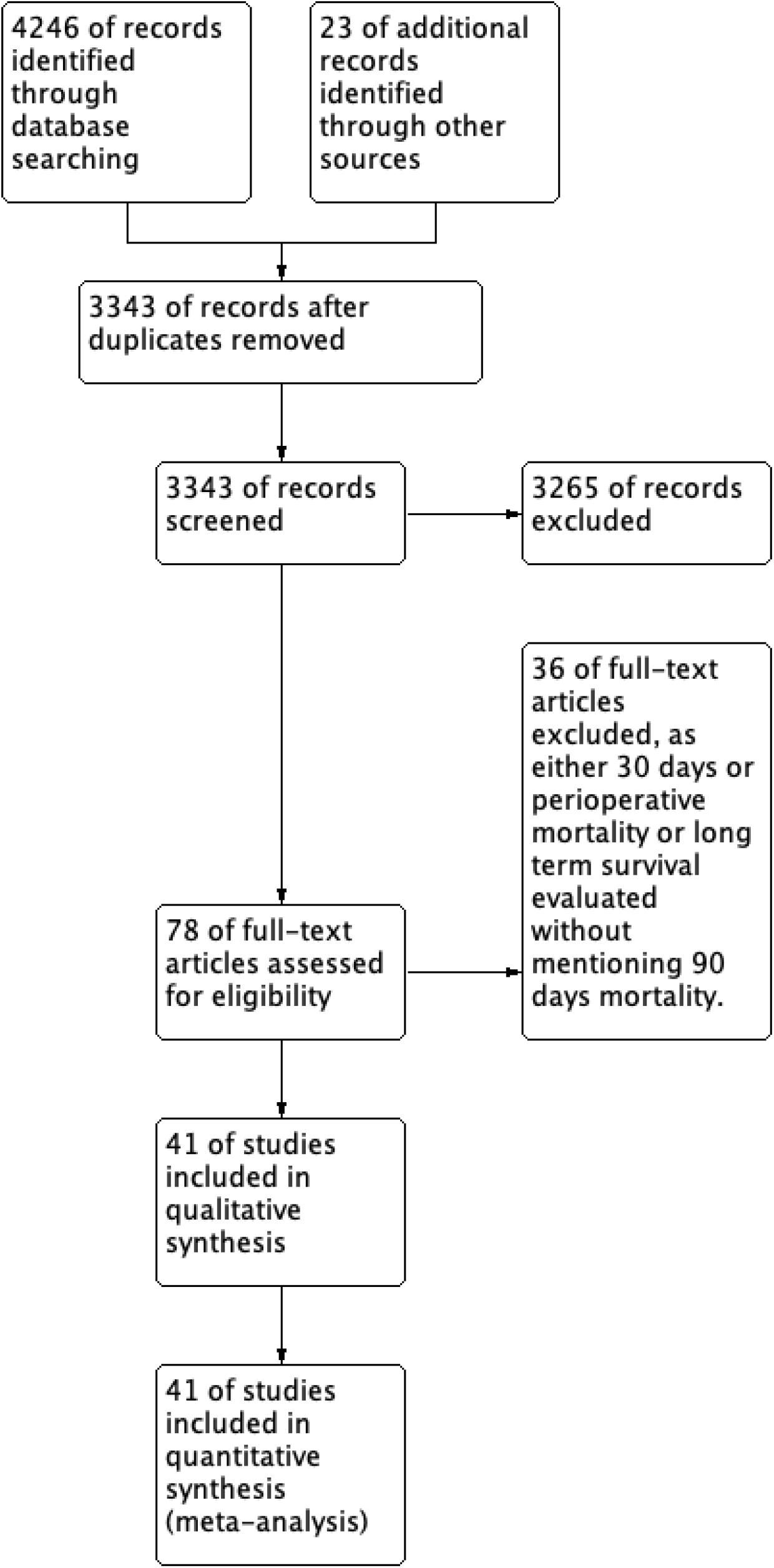
Prisma flow chart.

**Figure 2.**
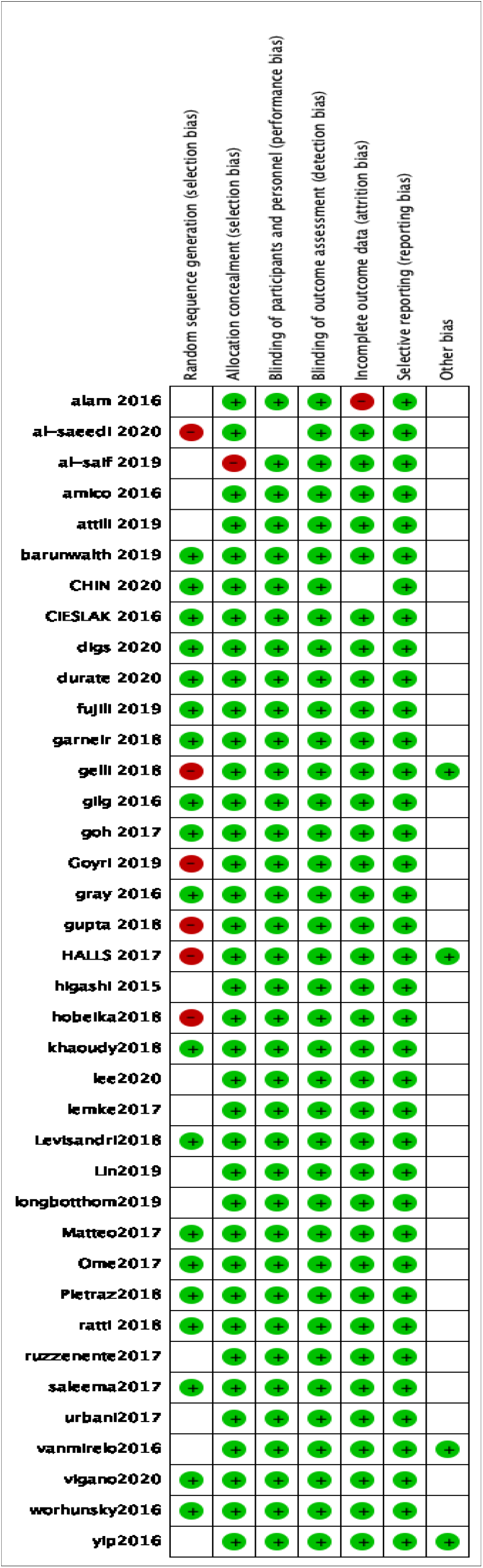
Risk of Bias summary

### Weighted 90 days mortality

Total 29931 patients’ data who underwent liver resections for various etiologies were pooled from 41 studied included.[13-52].1257 patients died within 90 days post operatively. Weighted 90 days mortality was 3.6% with 95% confidence interval between 2.8% to 4.4%. [Figure 3 Forest plot]. However, heterogeneity of the analysis was high with I^2^ 94.625%, tau^2^ 0.001, and Q value of 744.188. P value of heterogeneity was highly significant (p<0.001).

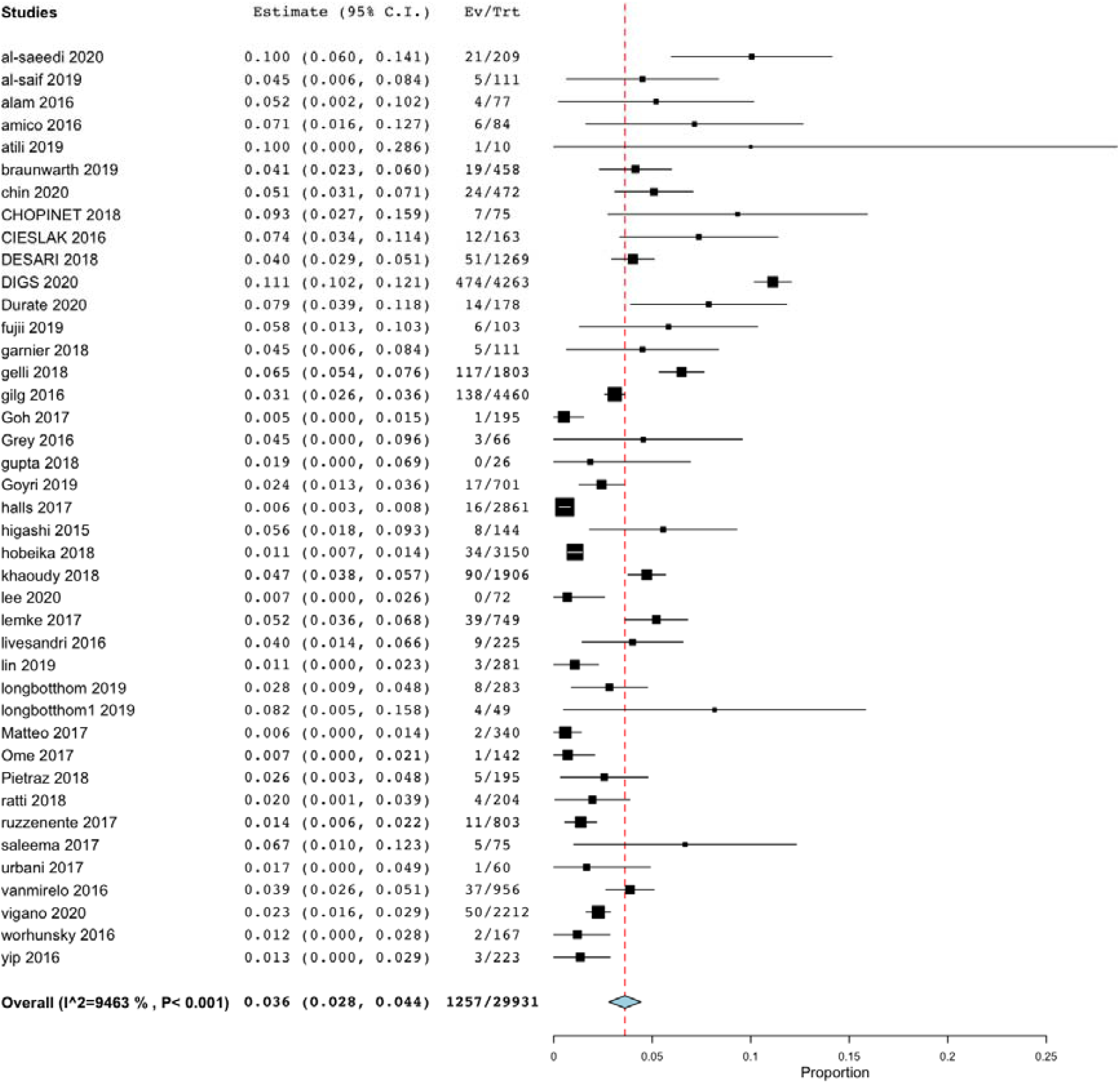

### Publication Bias

Figure 4 mentioned significant publication bias. Egger’s test showed significant publication bias with p value of <0.001.

**Figure 4.**
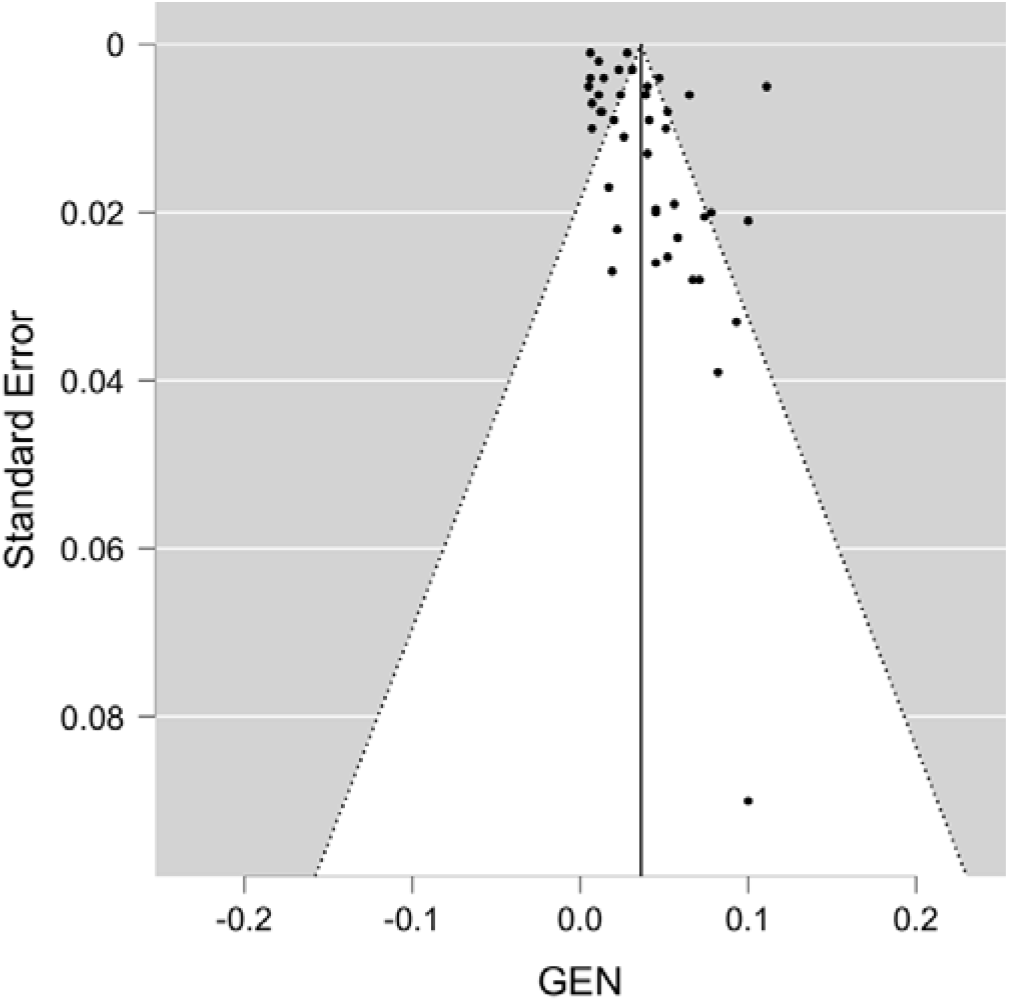
Funnel Plot for publication bias. (Egger’s test showed significant publication bias, p <0.001).

### Metaregression analysis

We analysed various covariates like major hepatectomy, Age of the patient, blood loss, open surgery, liver resections done for hepatocellular carcinoma or colorectal liver metastasis and cirrhotic liver to check for their association with heterogeneity in the analysis and hence 90 days mortality.

On univariate metaregression analysis major hepatectomy (p<0.001), Open hepatectomy (p<0.001), blood loss (p=0.002) were associated with heterogeneity in the analysis and 90 days mortality. On multivariate metaregression model after entering above 3 factors which were significant in univariate model, blood loss was significantly associated with heterogeneity and 90 days mortality. (p value= 0.03). However, residual heterogeneity was still highly significant with I^2^75.74% and p <0.001. So, we entered all the factors in multivariate model. On all covariates multivariate model, Major hepatectomy(p=0.003) and Open surgery (p=0.012) was independently associated with higher 90 days mortality, and liver resection for colorectal liver metastasis was independently associated with lesser 90 days mortality (z= −4.11,p<0.01). Residual heterogeneity after all factor multivariate metaregression model was none (I^2^=0,Tau^2^=0, H^2^=1) and nonsignificant (p=0.49) suggesting homogeneity of the findings. However only 10 studies mentioned all the factors. Metaregression forest plots for each parameters) is mentioned in supplement Figure 1 and multivariate forest plot (All Factors) mentioned in Figure 5. Funnel plot of studies mentioned all the factors and entered in multivariate model showed no publication bias. (p= 0.285 by eggers’ test). [Figure 6]. Normal Q-Q plot after meta-regression analysis is shown in Figure 7.

**Figure 5.**
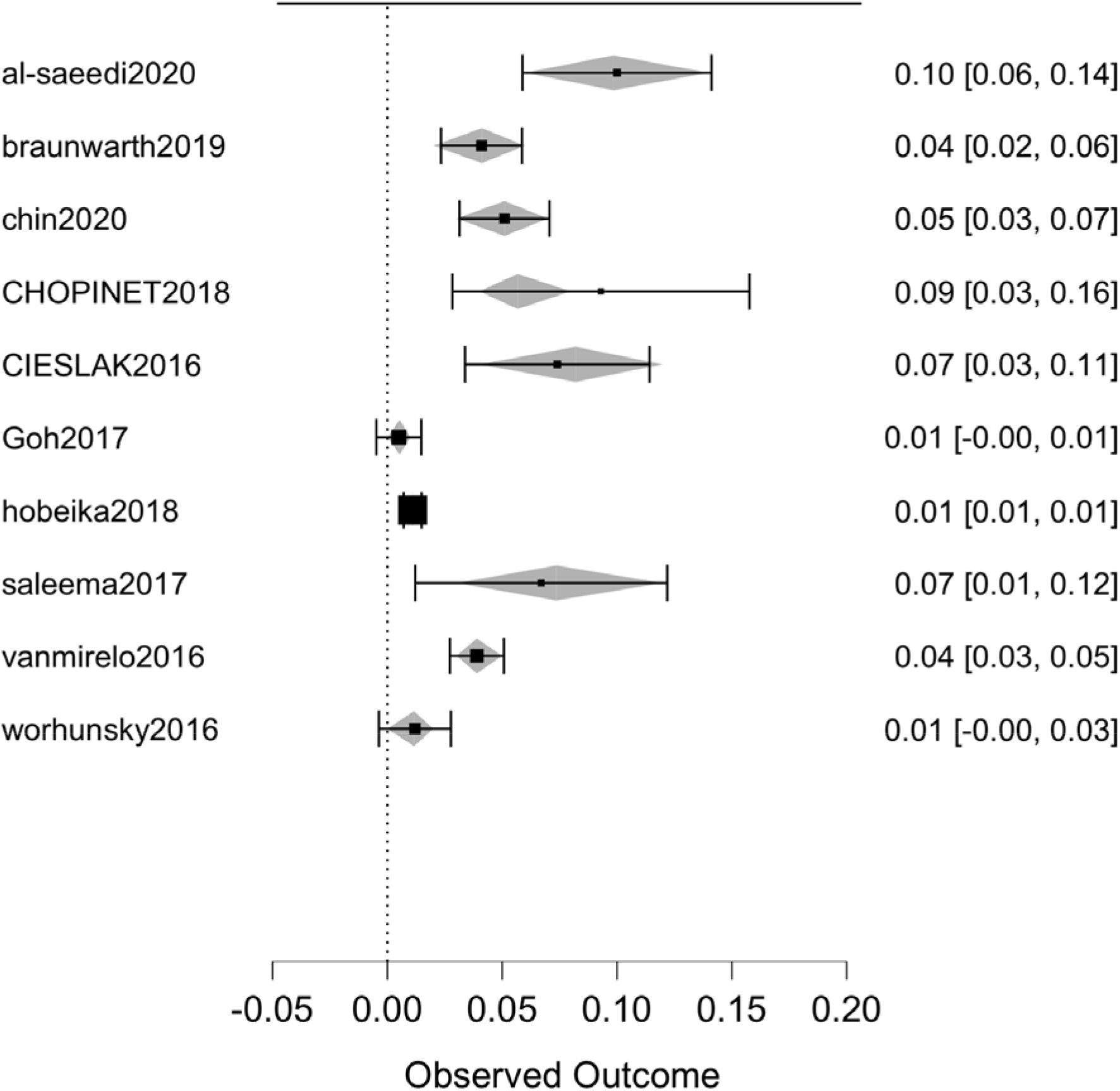
Forest plot multivariate metaregression. (grey squares is expected effect size with regard to covariate)

**Figure 6.**
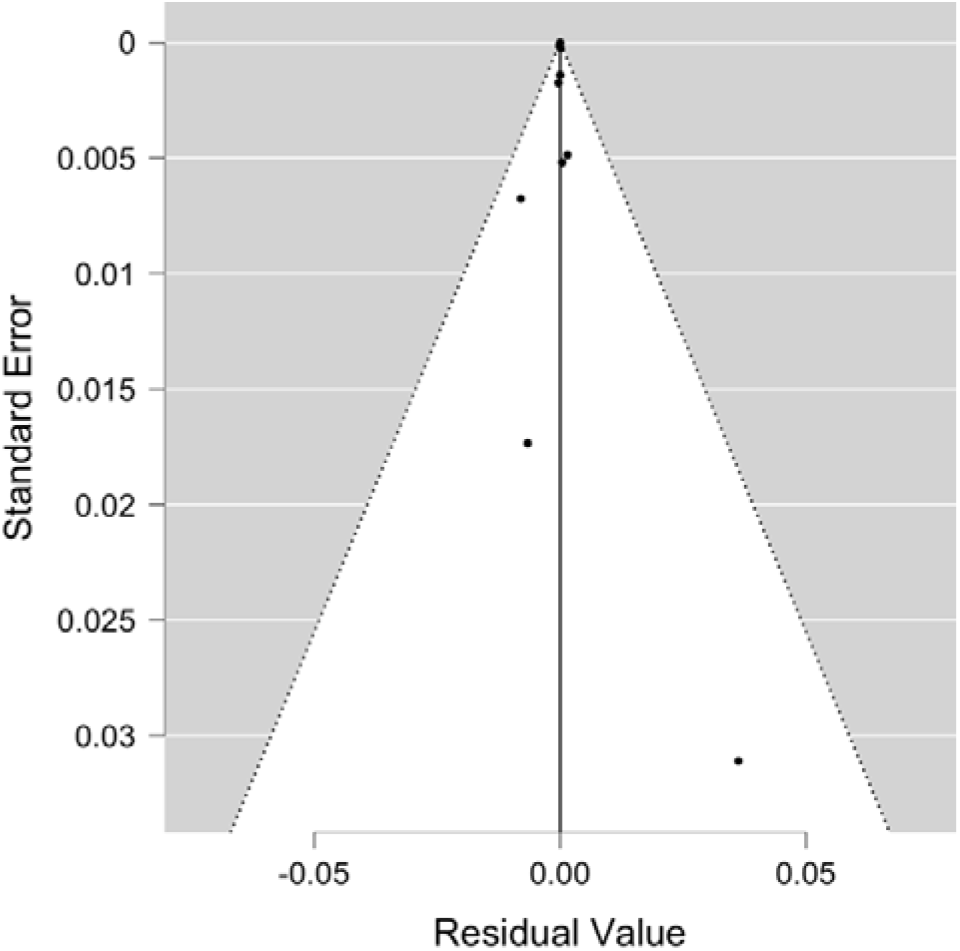
Funnel plot of publication bias for studies containing all the covariate in metaregression. (p=0.285 by egger’s test)

**Figure 7.**
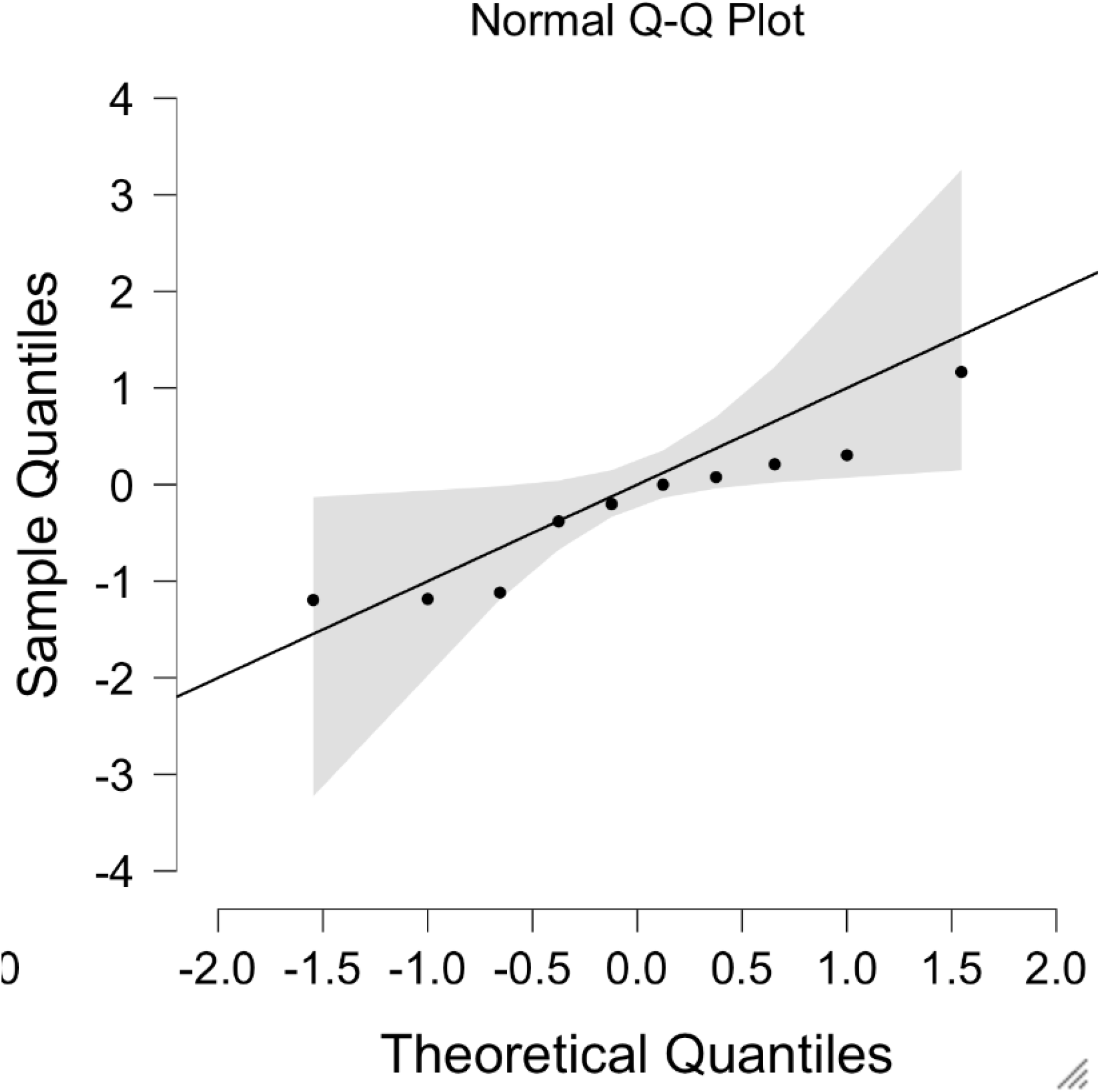
Normal Q-Q plot.

## Discussion

Liver resection is a curative surgery for many benign and malignant disorders, with most common indications are hepatocellular carcinoma and colorectal liver metastasis. Liver resection was historically associated with very high morbidity and mortality, which has now decreased significantly due to improved surgical and anaesthetic technique and improved perioperative and critical care. However, it is still considered to be associated with high mortality. Particularly recent study by Filmann et al [6],showed very high 30 days mortality after liver resection in Germany, they also raised concerns about possible selection and reporting bias in mortality studies.

There is also wide variation in defining peri/post-operative mortality in liver surgeries with some centres reporting in hospital mortality, some centres reporting 30 days and some centres reports 90 days mortality. Mayo et al [53] described that 90 days after surgery should be the standard criteria for defining peri/post-operative mortality. There is also wide regional variations in indications of liver resections. In Asian countries most common indication is hepatocellular carcinoma [54] while in European countries most common indications is colorectal metastasis, which can also be reason of variable mortality following liver resection. Our Aim in conducting this systemic review and prevalence meta-analysis to study weighted post-operative mortality rates after liver resections. We also aimed to look at the heterogeneity of the analysis and publication bias. We also did metaregression analysis for various factor affecting mortality like Major or minor hepatectomy, blood loss, age of the patient, open vs laparoscopic liver resections, cirrhotic background of the liver and etiologies for resections like hepatocellular carcinoma and colorectal metastasis in study published in last 5 year to look for recent trends. As we wanted to analyse 90 days mortality as standard definitions we excluded all the studies which did not mention 90 days mortality and excluded studies which studied only hepatocellular carcinoma or colorectal liver metastasis and included studies which analysed mortality rates in liver resections performed for various etiologies.

As shown in Figure 3 weighted 90 days mortality after liver resection was 3.6% with 95% confidence interval between 2.8%- 4.4%. However, heterogeneity was significantly high with I^2^=94.63% and p value <0.001. Suggesting various Moderators was responsible for this variable effect sizes in different studies. There was also significant publication bias as shown in funnel plot [Figure 4], and egger’s test was also significantly positive for publication bias.

Therefor we evaluated various factors affecting mortality like age, major hepatectomy, open surgery, blood loss, cirrhotic background, liver resection for hepatocellular carcinomas and colorectal liver metaanalysis.

In univariate metaregression analysis major hepatectomy, open surgery and blood loss was significantly associated with heterogeneity of analysis. When we initially performed metaregression analysis including factors those were significant in univariate analysis, residual heterogeneity of the analysis was still highly significant suggesting various other factors or mixed effect of various factors on mortality was responsible for heterogeneity. So to evaluate that we entered all the factors available in the metaregression model. That showed no residual heterogeneity with I^2^=0, Tau^2^=0 and H^2^ =1 with nonsignificant p value of the heterogeneity (p=0.49) suggesting mixed effect of all the factors was responsible for variable effect size and mortality in studies. In multivariate metaregression with all the moderators open hepatectomy (p=0.012), Major hepatectomy (p=0.003), and cirrhotic liver (p=0.003) was independently associated with higher mortality and colorectal metastasis was independently associated with lower 90 days mortality. (p<0.0001, z = −4.116). Elimination of residual heterogeneity after metaregression suggested that above factors was mainly responsible for variable outcomes across the centres. Q-Q plot also suggested lack of heterogeneity after metaregression. [Figure 7].

Limitations of the meta-analysis were some large number studies had to be excluded due to lack of 90 days mortality data for example filmann et al [6] study which evaluated 30 days mortality rate in Germany. Also, we could not take in account centre volume and surgeon’s experience. Another limitations was only 10 studies mentioned all the moderators however, number of study was adequate to conduct metaregression analysis.

However, to our knowledge this is the first meta-analysis evaluated weighted 90 days mortality rates and evaluated various factors responsible for heterogeneity and lack of residual heterogeneity after metaregression proved their effects on variable mortality rates across the centres.

In conclusion reporting of perioperative mortality rates should be standardised as 90 days mortality rates and, they should be evaluated in context of various moderators described above. Major hepatectomy, open hepatectomy, and cirrhotic background is associated with higher mortality rates and colorectal liver metastasis is associated with lower peri operative mortality rates compared to other etiologies like hepatocellular carcinoma.

## Data Availability

data will be made available on demand.

## Notes

Conflict of Interests: None

Funding disclosure: nothing to disclose.

### Competing Interest Statement

The authors have declared no competing interest.

### Funding Statement

no funding disclosure

### Author Declarations

Shalby hospital IRB board.

