## supplement file 1 for "Recent trends in postoperative mortality after liver resection- A systemic review and metanalysis of studies published in last 5 years and metaregression of various factors affecting 90 days mortality"

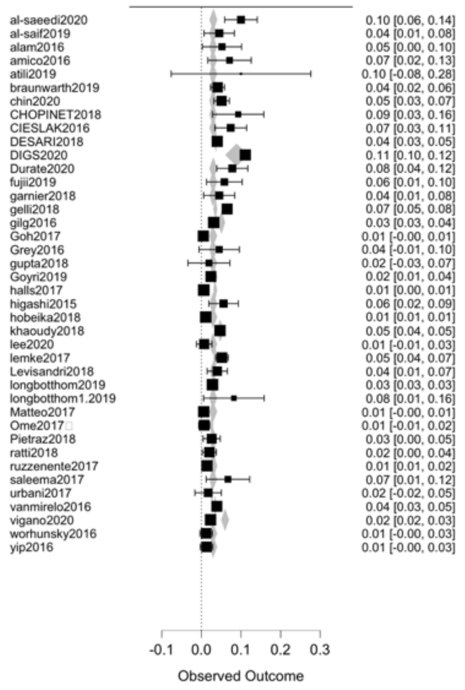


Supplement Figure 1(a): metaregression forest plot for major hepatectomy

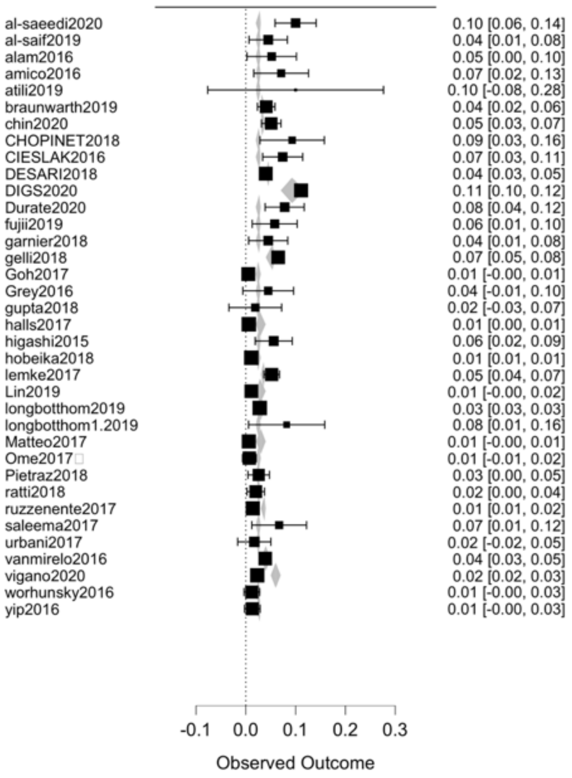


Supplement Figure 1(b): metaregression forest plot for open hepatectomy.

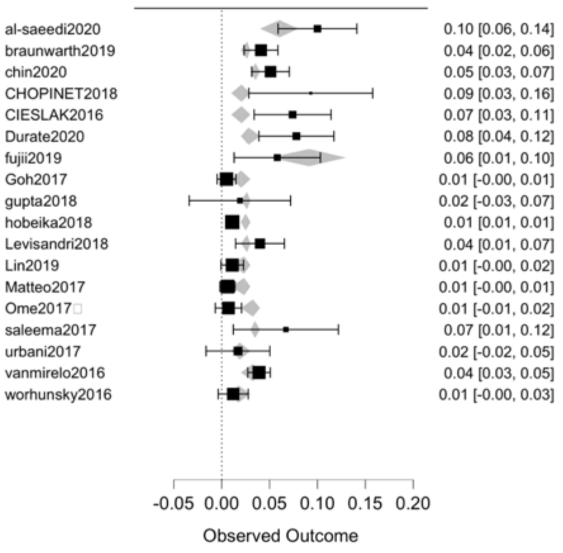


Supplement Figure 1 (c) metaregression forest plot for blood loss.

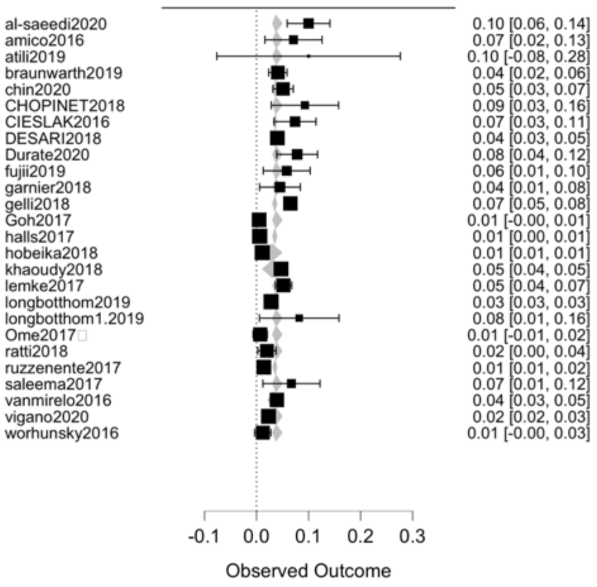


Supplement Figure 1(d): metaregression forest plot for cirrhosis.

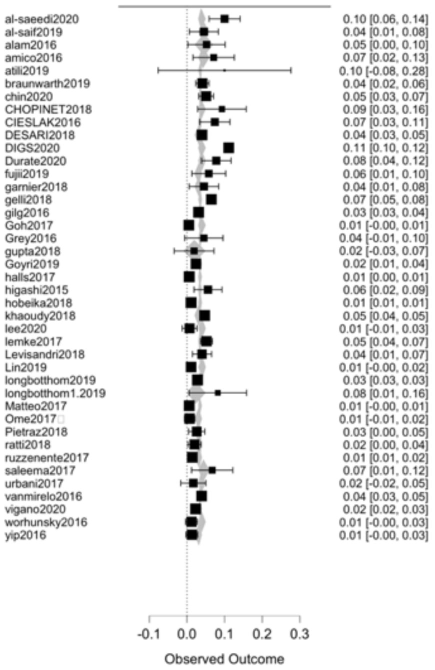


Supplement Figure 1(e) metaregression forest plot for Age.

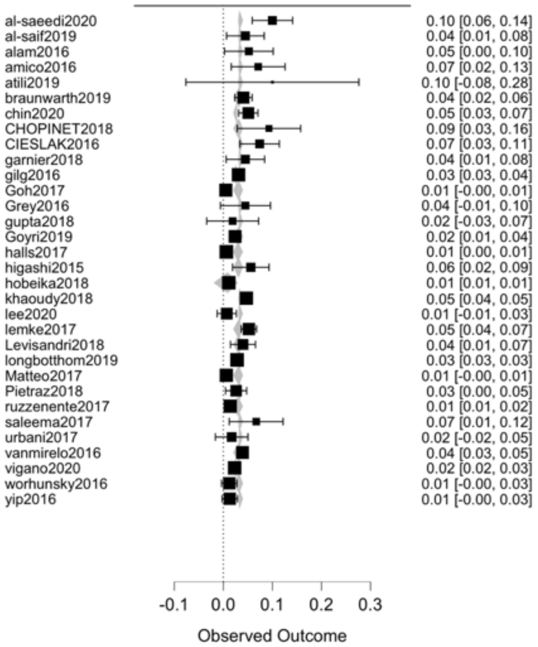


Supplement Figure 1(f): metaregression forest plot for Hepatocellular carcinoma.

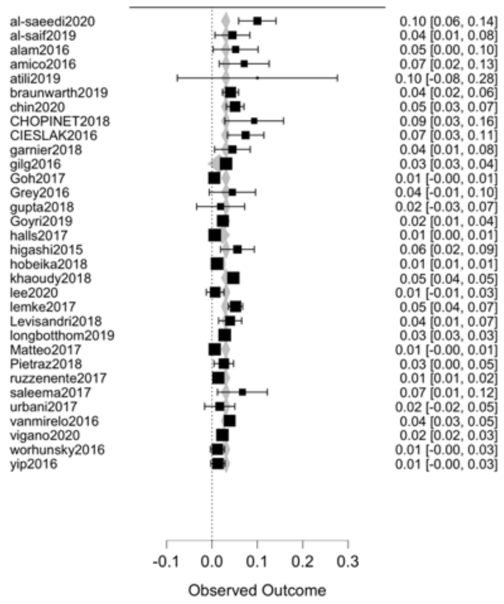


Supplement Figure 1(g): meta-regression Forrest plot for Colorectal liver metastasis
